# Cell-type-specific polygenic risk scores reveal adipocyte-related interactions with lipids in coronary artery disease

**DOI:** 10.64898/2026.07.07.26357510

**Authors:** Jiaqi Hu, Leqi Xu, Tianyu Liu, Wangjie Zheng, Hongyu Zhao

## Abstract

**Background:** Genome-wide polygenic risk scores (PRSs) for coronary artery disease (CAD) aggregate genetic effects across the genome and may obscure biologically distinct mechanisms. We aimed to develop cell-type-specific PRSs (csPRSs) using single-cell RNA sequencing (scRNA-seq) data and investigate their interactions with lipids on CAD risk.

**Methods:** Using publicly available scRNA-seq data from human heart tissue, we identified cell-type-specific genes across 13 major cell types and 64 subpopulations and grouped them into 10 cell clusters. Variants from a CAD genome-wide association study (GWAS) were mapped to cluster-specific genes to construct csPRSs for European-ancestry participants from the UK Biobank (UKB). Interactions between csPRSs and lipid-related phenotypes were evaluated using Cox proportional hazards models and stratified analyses, with significant findings further assessed in an internal validation dataset.

**Results:** Distinct interaction patterns with lipid phenotypes were observed across csPRSs. Low-density lipoprotein (LDL)-related lipid traits, including apolipoprotein B (ApoB), low-density lipoprotein cholesterol (LDL-C), and total cholesterol (cholesterol), primarily interacted with adipocytes (Adip), whereas high-density lipoprotein (HDL) traits interacted with endothelial-mesothelial (EC-Meso), fibroblast (FB), and immune-cell csPRSs. Notably, interactions for Adip csPRSs were replicated in internal validation analyses.

**Conclusions:** Cell-type-specific decomposition of genome-wide PRSs for CAD identified biologically distinct lipid interactions that were not captured by the genome-wide PRS. Adipocyte genetic factors may influence how LDL lipids affect CAD risk. These findings highlight the potential of cell-type-informed PRSs to improve the biological interpretation of PRSs and provide insights into the heterogeneous mechanisms underlying CAD.

## Introduction

Coronary artery disease (CAD) is a leading cause of morbidity and mortality worldwide^1^. Twin studies and large-scale genome-wide association studies (GWASs) have demonstrated substantial heritability for CAD^2,3^, supporting the use of polygenic risk scores (PRSs) for risk stratification. Multiple CAD PRSs have shown robust predictive performance across populations^4–7^. In addition, prior studies identified significant interactions between CAD PRSs and lipid traits, particularly low-density lipoprotein cholesterol (LDL-C)^8^. Specifically, LDL-C was more strongly associated with CAD in individuals at high genetic risk for CAD than in those at intermediate or low genetic^8^, with implications for precision prevention. However, genome-wide PRSs aggregate variants across the entire genome into a single score, limiting biological interpretability and potentially obscuring heterogeneous disease mechanisms underlying the interactions^9^.

To improve biological resolution, several approaches have been developed to decompose genome-wide PRSs into biologically informed components, e.g., pleiotropy-decomposed PRS (PD-PRS), which partitioned CAD PRSs into pleiotropy-informed components, including lipid-related PD-PRSs^10^. Although lipid-related PD-PRSs showed strong associations and interactions with lipid traits, the underlying cellular mechanisms remained unclear. More recently, an endothelial cell (EC)-specific PRS was reported to significantly interact with LDL-C on CAD risk^11^, suggesting that gene-lipid interactions may involve cell-type-specific pathways. However, restricting analyses to EC-related genes may overlook the contributions of other relevant cell types, including adipocytes, fibroblasts, and immune cells. In addition, variants assigned to one cell type may function across multiple cell populations. For example, rs1879454 near *MESDC1*, included in the EC-PRS, has also been implicated in fatty acid uptake in CD8+ T cells^12^. These observations suggest that a more comprehensive decomposition of PRSs across multiple cardiac and vascular cell types may better capture the biological mechanisms underlying gene-lipid interactions in CAD.

In this study, we proposed a framework to construct cell-type-specific PRSs (csPRSs) using published single-cell RNA sequencing (scRNA-seq) data from human heart tissue and investigated their interactions with lipid traits on CAD risk. We grouped major cardiac and vascular cell populations into 10 clusters and constructed csPRSs using variants mapped to cluster-specific genes. Among unrelated participants of European (EUR) ancestry from the UK Biobank (UKB), csPRSs demonstrated distinct interaction patterns with lipid traits. In particular, apolipoprotein B (apoB), LDL-C, and total cholesterol (cholesterol) showed significantly stronger associations with CAD among individuals with higher adipocyte csPRSs. These findings were further investigated in an internal validation cohort, highlighting the value of cell-type-informed PRSs for understanding biological heterogeneity in CAD.

## Methods

### Overview of Methods

The study overview is summarized in Figure 1. Using publicly available scRNA-seq data from human heart tissue^13^, we identified cell-type-specific genes across 13 major cell types and 64 subpopulations through hierarchical clustering. Based on similarities in gene expression profiles, these cell types were grouped into 10 clusters. Variants from a large-scale GWAS for CAD were mapped to genes by genomic position to define cell-cluster-specific variant sets for csPRS construction. Unrelated EUR participants from the UKB were divided into training, testing, and validation datasets. In the training dataset, we compared two strategies for constructing csPRSs and selected the optimal approach for downstream analyses. In the testing dataset, we evaluated associations of csPRSs with incident CAD and major CAD risk factors, and tested interactions between csPRSs and seven lipid traits as well as lipid-lowering medication use. Significant interactions were subsequently replicated in the internal validation dataset.

**Figure 1.**
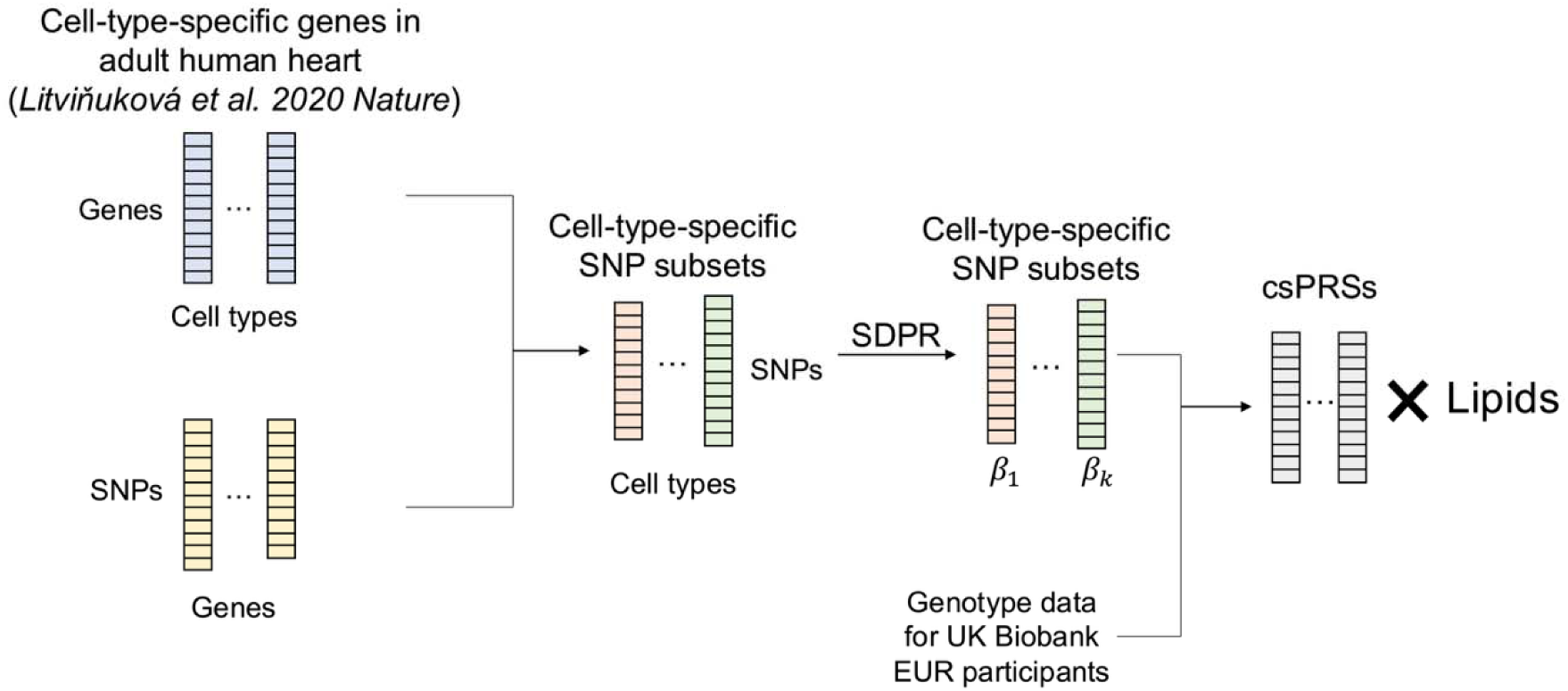
Study overview. Overview of the study. Publicly available scRNA-seq data from human heart tissue were used to identify cell-type-specific genes across 13 major cell types and 64 subpopulations, which were further grouped into 10 cell clusters. Variants from a large-scale CAD GWAS were mapped to cluster-specific genes to construct csPRSs. Unrelated EUR participants from the UKB were divided into training, testing, and validation datasets. In the training dataset, two csPRS construction strategies were compared and the optimal approach was selected. Associations of csPRSs with incident CAD, CAD risk factors, and interactions with lipid traits and lipid-lowering medication use were evaluated in the testing dataset and replicated in the internal validation dataset.

### Study samples

Individual-level genotype and phenotype data from the UKB were used in this study. UKB is a large prospective cohort study that enrolled over 500,000 participants aged 40-69 years across the United Kingdom^14^. CAD was defined using self-reported diagnoses and linked hospital records based on International Classification of Diseases, Ninth and Tenth Revisions (ICD-9 and ICD-10), and Office for National Statistics Classification of Interventions and Procedures (OPCS) codes (Supplementary Table 1). Participants with any corresponding code were classified as CAD cases, whereas participants without a recorded CAD diagnosis were considered controls. To focus on incident CAD, individuals with prevalent CAD diagnosed before recruitment or with missing diagnosis dates were excluded. All analyses were restricted to unrelated participants of genetically inferred EUR ancestry^5^.

Phase III genotype data from the UKB were used in this study. Participants were genotyped using either the UK BiLEVE Axiom Array or the UK Biobank Axiom Array, covering approximately 820,000 variants. Genotypes were centrally imputed using the 1000 Genomes Project and Haplotype Reference Consortium (HRC) reference panels, resulting in approximately 93 million variants per individual. Standard quality control procedures were applied, retaining autosomal variants with an imputation quality score > 0.3, a Hardy-Weinberg equilibrium p-value > 1×10^-5^, and an MAF > 0.05.

Participants were categorized into Caucasian and non-Caucasian EUR groups based on UKB Field ID 22006 (genetic ethnic grouping). The Caucasian EUR group was randomly divided into training (60%) and testing (40%) datasets while the non-Caucasian EUR group was reserved for internal validation analyses.

### Cell type-specific gene sets

We used publicly available scRNA-seq data from the human heart^13^ to identify cell-type-specific genes as previously described^15^. In addition to the 13 major cell types, we also included 64 corresponding subpopulations to account for heterogeneity of gene expression in major cell types. Specifically, first, genes expressed in more than 1% of cells were retained, resulting in 14,204 genes for downstream analyses. Gene specificity (*Sg*) for gene *i* in cell type *j* was calculated as 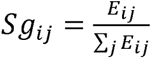, where *E_ij_* was the average expression of gene *i* in cell type *j*. For each cell type, genes were ranked according to specificity scores, and the top 10% were defined as cell-type-specific genes.

Because cell subpopulations were defined based on broader major cell types, overlaps among cell-type-specific gene sets were expected. To account for similarities across gene sets, we calculated pairwise Jaccard similarity, defined as the number of shared genes divided by the total number of unique genes between two sets. Hierarchical clustering was then performed using the pairwise Jaccard similarity matrix, with the number of clusters ranging from 2 to 20. The optimal number of clusters was selected based on the proportion of variance explained, overall similarity patterns, and biological knowledge.

For each cell cluster, functional enrichment was evaluated using overrepresentation analysis of KEGG pathways. For each cell cluster-pathway pair, a two-by-two contingency table was constructed based on whether genes were cell-cluster-specific and whether they belonged to the KEGG pathway of interest. Enrichment was assessed using Chi-squared tests under the null hypothesis that cell-cluster-specific genes were not overrepresented in the pathway. Pathways with false discovery rate (FDR)-adjusted p-values < 0.05 were considered significantly enriched.

### Construction of cell-type-specific PRSs (csPRSs)

A large-scale meta-analyzed GWAS summary statistics for CAD from individuals of EUR ancestry were used for PRS constructio^16^. The GWAS included 60,801 CAD cases and 123,503 controls, without inclusion of UKB as an individual study. Indels and duplicated variants were excluded, and analyses were restricted to HapMap3 variants present in the UKB genotype data to ensure high-quality variants for downstream PRS analyses.

Variants that passed quality control were mapped to genes based on their genomic positions. Specifically, variants located within 10 kilobases (kb) upstream or downstream of a gene were assigned to that gene. Cell-cluster-specific variant sets were then generated using genes specifically expressed within each cell cluster.

Two strategies were evaluated for csPRS construction using Summary statistics based Dirichlet Process Regression (SDPR)^17^. In the first strategy, SDPR was applied to the full GWAS summary statistics, and variants belonging to each cell cluster were subsequently grouped to generate csPRSs (genome-wide training). In the second strategy, SDPR was applied separately to each cell-cluster-specific variant set to directly estimate csPRS weights (cell-cluster training). We hypothesized that restricting training to biologically relevant variant sets could improve the specificity and predictive performance of csPRSs compared with genome-wide training.

The genome-wide PRS for CAD, generated using SDPR, and csPRSs trained with two strategies were calculated for UKB participants as weighted sums of risk alleles using PLINK 2.0^18^. All PRSs were standardized to mean 0 and standard deviation 1 within the training, testing, and validation datasets, separately.

### Statistical analyses

#### Associations between PRSs and incident CAD

Associations between PRSs and incident CAD were evaluated using Cox proportional hazards models. Follow-up time was defined as the interval from recruitment to CAD diagnosis, loss to follow-up, death, or end of follow-up, whichever occurred first. Hazard ratios (HRs) per standard deviation increase in PRS were estimated, and predictive performance was assessed using the Harrell’s concordance index (C-index). To evaluate whether cell-type-specific weight training improved CAD risk prediction, HRs and C-indices calculated in training dataset were compared between the two csPRS training strategies. The strategy with better predictive performance was selected for downstream analyses.

We further evaluated whether integrating csPRSs and their corresponding complement csPRSs (c-csPRSs) could improve CAD risk prediction compared with genome-wide PRSs alone. The c-csPRSs were constructed using variants not included in each corresponding csPRS, with weights estimated using SDPR. Prediction models for incident CAD were developed in the training dataset using csPRSs and c-csPRSs as predictors within Cox proportional hazards frameworks. Three modeling approaches were evaluated: ridge regression^19^ and elastic net regression^20^ implemented using R package glmnet^21^, and XGBoost^22^ implemented using R package xgboost. Prediction accuracy was evaluated in the testing samples using C-index.

#### Associations between csPRSs and CAD risk factors

To explore potential biological mechanisms linking csPRSs to CAD risk, we evaluated associations between csPRSs and 71 CAD-related risk factors in the testing dataset. These phenotypes were grouped into 18 categories, including alcohol consumption, asthma, autoimmune disease, blood pressure, body measurements, cancer, cardiovascular and atherosclerotic diseases, diabetes, immune function, kidney function, lipids, liver enzymes, metabolic factors, neuropsychiatric diseases, osteoporosis, protein biomarkers, sex hormones, and smoking. Associations were assessed using logistic regression for binary traits and linear regression for continuous traits.

#### Interactions between csPRSs and lipid phenotypes

Interactions between csPRSs and eight lipid-related phenotypes were evaluated, including lipid-lowering medication use (med), triglycerides (TG), lipoprotein(a) (LipoA), cholesterol, LDL-C, ApoB, high-density lipoprotein cholesterol (HDL-C), and apolipoprotein A (ApoA). For analyses involving lipid measurements other than medication use, participants taking lipid-lowering medications were excluded.

First, multiplicative interactions between csPRSs and lipid phenotypes were analyzed using the Cox proportional hazards model. This analysis provided an overall assessment of statistical interactions between genetic risk and lipid-related phenotypes on incident CAD risk. However, interpretation of interaction effects was limited by differences in phenotype scales. Therefore, we additionally performed stratified analyses using binary lipid phenotypes and tertiles of csPRSs to facilitate clinical interpretation.

For improved interpretability, the seven continuous lipid measurements were dichotomized according to current clinical guidelines (Supplementary Table 2). The csPRSs were categorized into low, intermediate, and high groups based on tertiles. Associations between binary lipid phenotypes and incident CAD within each csPRS tertile were evaluated using Cox proportional hazards models, and interactions were tested by modeling csPRS tertiles as ordinal variables. Models were adjusted for age at recruitment, sex, and the top 10 genetic principal components. Absolute interactions were additionally assessed using absolute risk (AR), defined as the proportion of participants developing incident CAD within each lipid-csPRS subgroup. Absolute risk reduction (ARR) was calculated as the difference in AR between harmful and normal lipid groups. Trends in ARR across csPRS tertiles were evaluated by testing the interaction between ordinal csPRS tertiles and binary lipid phenotypes without covariate adjustment.

All interaction analyses were performed in the testing dataset. Lipid-csPRS pairs showing at least nominal significance (P<0.05) in tertile-based interaction analyses were retained for downstream validation.

#### Characteristics of participants with high csPRSs

We further hypothesized that CAD patients with different csPRS levels may exhibit distinct disease trajectories before CAD onset. To characterize pre-CAD disease patterns, we used previously developed disease embeddings, in which each ICD-10 code is represented by encoding its associated clinical description using a GPT-based embedding mode^7,23^. CAD patients with diagnoses of atrial fibrillation (AF), angina, chronic kidney disease (CKD), heart failure (HF), ischemic stroke (IS), peripheral artery disease (PAD), and type 2 diabetes (T2D) before CAD onset were included. Individual-level embeddings were calculated as the average embedding corresponding to diseases diagnosed before CAD onset. Differences in disease trajectory patterns between individuals within and outside the top csPRS tertile were evaluated using permutational multivariate analysis of variance (PERMANOVA).

All statistical analyses were performed using R version 4.2.0. Unless otherwise specified, statistical significance was defined as P<0.05.

## Results

### Study samples

A total of 308,072 unrelated EUR participants without prevalent CAD at recruitment from the UKB were included in the analyses, among whom 13,623 incident CAD cases were identified during follow-up. Baseline characteristics are summarized in Table 1. Compared with non-CAD controls, participants who developed incident CAD were older at recruitment, more likely to be male and ever smokers, had higher body mass index (BMI), and showed less favorable lipid profiles. The unrelated Caucasian EUR population was further divided randomly into training (7,184 cases and 153,457 controls) and testing (4,779 cases and 102,315 controls) datasets. The internal validation was conducted using non-Caucasian EUR (1,660 cases and 38,677 controls).

**Table 1.** Demographic characteristics.

|  | CAD cases<br>(N = 13,623) | Controls<br>(N = 294,449) | P |
| --- | --- | --- | --- |
| Age at recruitment | 60.09 (6.92)* | 56.28 (8.03) | <2E-16 |
| Sex (Male) | 9,438 (69.3%) | 129,873 (44.1%) | <2E-16 |
| BMI (Kg/m<sup>2</sup>) | 28.58 (4.77) | 27.22 (4.75) | <2E-16 |
| Ever smoking | 7,576 (55.9%) | 131,270 (44.8%) | <2E-16 |
| ApoA (g/L) | 1.46 (0.25) | 1.56 (0.27) | <2E-16 |
| ApoB (g/L) | 1.16 (0.24) | 1.06 (0.23) | <2E-16 |
| Cholesterol (mmol/L) | 6.15 (1.10) | 5.90 (1.06) | <2E-16 |
| HDL-C (mmol/L) | 1.33 (0.34) | 1.49 (0.38) | <2E-16 |
| LDL-C (mmol/L) | 3.99 (0.84) | 3.71 (0.81) | <2E-16 |
| LipoA (nmol/L) | 51.39 (53.00) | 43.12 (48.40) | <2E-16 |
| TG (mmol/L) | 2.09 (1.15) | 1.68 (0.99) | <2E-16 |
| Med | 3,848 (28.2%) | 41,116 (14.0%) | <2E-16 |

\*: mean (standard deviation) for continuous variable and count (proportion) for binary variable.

### Development of csPRSs

Using scRNA-seq data from human heart tissue, we generated cell-type-specific gene sets across 13 major cell types and 64 subpopulations. The number of genes per cell type ranged from 686 for nucleus pulposus (NP) cells to 1,421 for EC (Supplementary Table 3). Hierarchical clustering based on pairwise Jaccard similarity of gene sets was then performed, evaluating cluster numbers from 2 to 20. Based on the proportion of variance explained and biological interpretability (Supplementary Figure 1), we selected nine clusters and further separated atrial and ventricular cardiomyocytes, resulting in 10 final cell clusters: atrial cardiomyocyte (aCM), ventricular cardiomyocyte (vCM), adipocyte (Adip), Immune 1, Immune 2, endothelial cell (EC), endothelial-mesothelial (EC-Meso), fibroblast (FB), neuronal cell (NC), and pericyte-smooth muscle cell (PC-SMC) (Figure 2).

**Figure 2.**
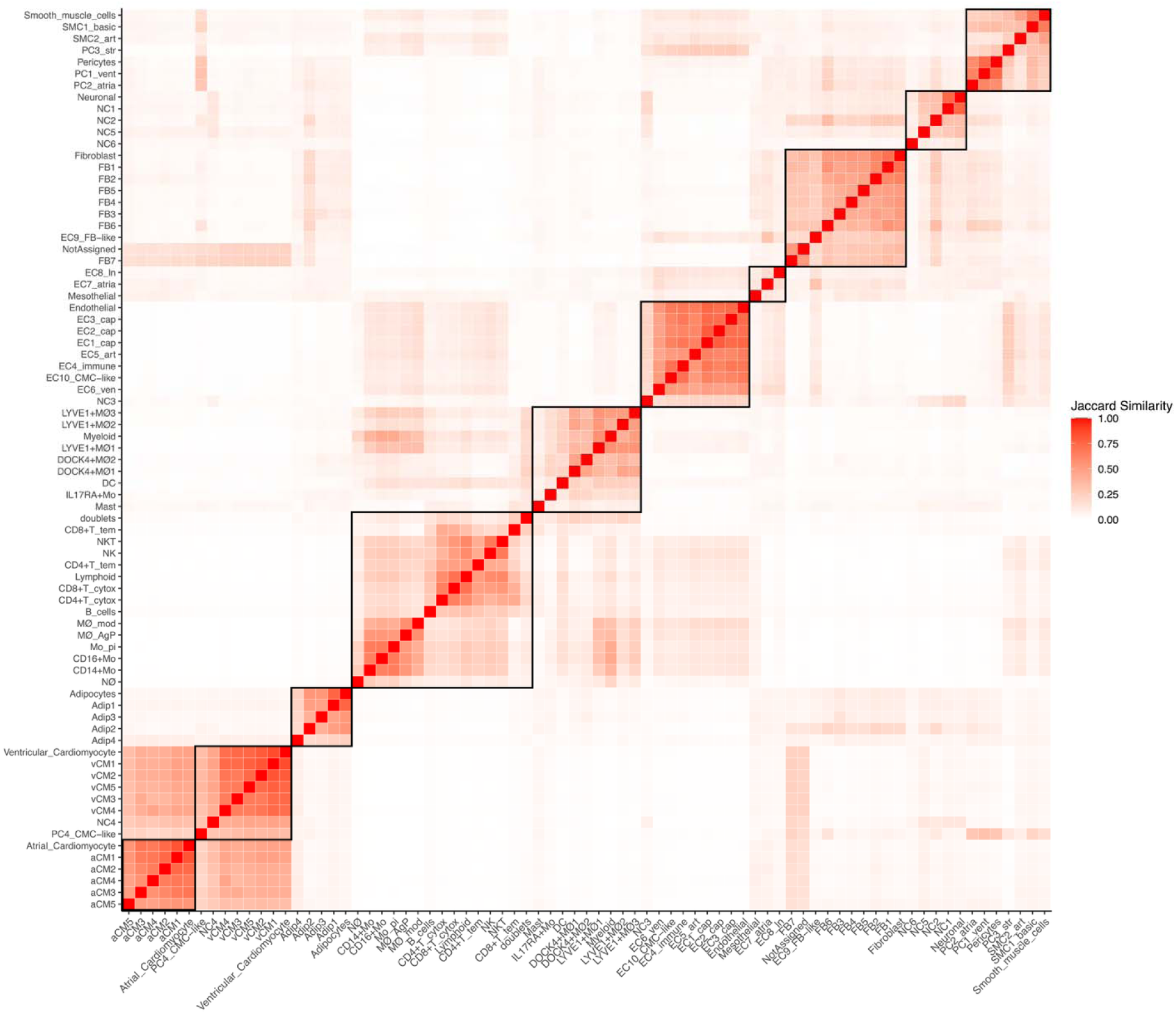
Clustered Jaccard similarity across cell-type-specific gene lists. We calculated the pairwise Jaccard similarity across gene lists for 13 major cell types and 64 subpopulations and clustered them. Each cell represented the Jaccard similarity between two gene lists, and the final 10 clusters were highlighted using black rectangles: aCM, vCM, Adip, Immune 1, Immune 2, EC, EC-Meso, FB, NC, PC-SMC.

Most clusters consisted of a major cell type and related subpopulations. For example, the aCM cluster included atrial cardiomyocyte and five related subpopulations. Notably, endothelial-related clusters separated EC into EC, EC-Meso, and FB clusters, suggesting substantial heterogeneity in gene-expression patterns across endothelial-associated subpopulations. Overall, similarities between clusters were modest, with most pairwise Jaccard similarities below 20% (Supplementary Figure 2).

Pathway overrepresentation analyses demonstrated significant enrichment of biologically relevant KEGG pathways within corresponding clusters (Supplementary Table 4). For example, the Adip cluster was enriched for pathways related to peroxisome proliferator-activated receptor (PPAR) signaling, regulation of lipolysis in adipocytes, and fatty acid metabolism. These results supported the biological validity of our inferred cell clusters.

Next, genes were mapped to variants included in the CAD GWAS summary statistics based on genomic position to construct 10 cell-cluster-specific variant sets. Partitioned heritability for each set was estimated using LDSC^25,26^. Among the 10 clusters, the PC-SMC set showed significant heritability enrichment, accounting for 5.52% of the total CAD heritability (P < 0.05) (Supplementary Table 5).

Two csPRS training strategies, genome-wide and cell-cluster training, were evaluated across the 10 cell-cluster-specific variant sets. The csPRSs generated from each strategy were calculated in the UKB and compared for CAD risk prediction using HRs and C-indices in the training dataset. Across all 10 clusters, csPRSs derived from the cell-cluster training strategy showed consistently higher HRs than those derived from genome-wide training (Figure 3, Table 2, and Supplementary Table 6). Relative improvements in HRs ranged from 1.70% for aCM to 4.85% for Immune2. Cell-cluster training yielded slight improvements in C-index, and the overall predictive performance remained moderate. These results suggest that incorporating cell-type-specific information during PRS training may improve association strength.

**Figure 3.**
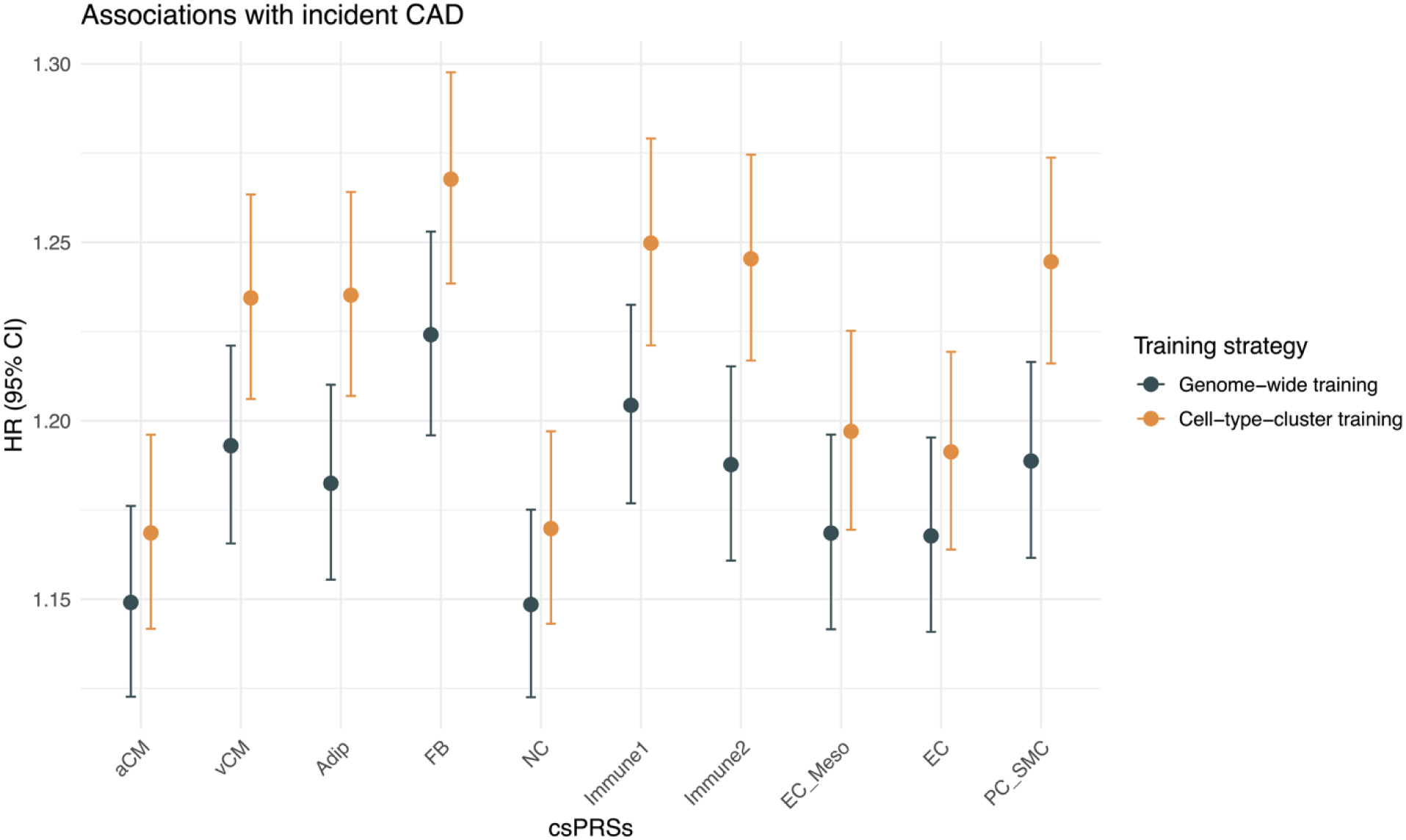
HR of csPRSs trained by two strategies. Associations between csPRSs trained by genome-wide (black) and cell-type-cluster (gold) strategies and incident CAD among UKB training dataset. HRs and 95% CIs were presented on y-axis across 10 csPRSs on the x-axis. Cell-type-cluster strategy showed higher HRs across all ten csPRSs.

**Table 2.**
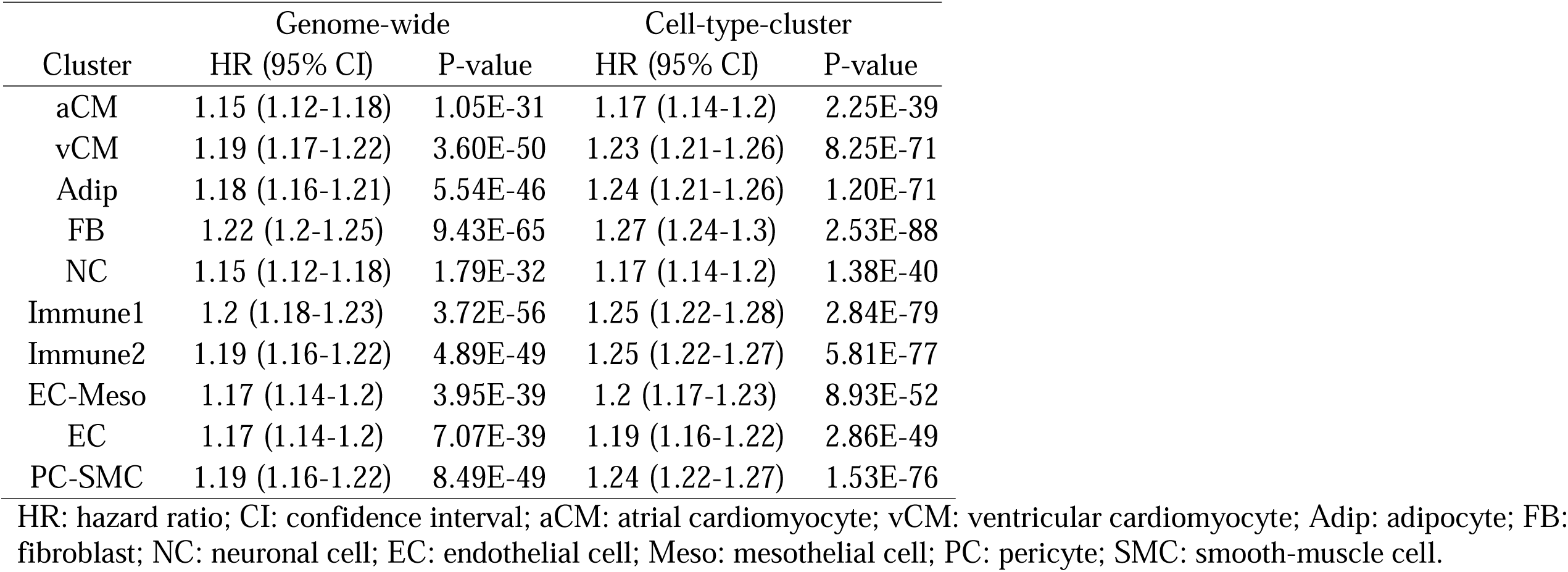
Hazards ratios (HRs) for csPRSs trained using genome-wide and cell-cluster-specific strategies.

| Cluster | Genome-wide |  | Cell-type-cluster |  |
| --- | --- | --- | --- | --- |
|  | HR (95% CI) | P-value | HR (95% CI) | P-value |
| aCM | 1.15 (1.12-1.18) | 1.05E-31 | 1.17 (1.14-1.2) | 2.25E-39 |
| vCM | 1.19 (1.17-1.22) | 3.60E-50 | 1.23 (1.21-1.26) | 8.25E-71 |
| Adip | 1.18 (1.16-1.21) | 5.54E-46 | 1.24 (1.21-1.26) | 1.20E-71 |
| FB | 1.22 (1.2-1.25) | 9.43E-65 | 1.27 (1.24-1.3) | 2.53E-88 |
| NC | 1.15 (1.12-1.18) | 1.79E-32 | 1.17 (1.14-1.2) | 1.38E-40 |
| Immune1 | 1.2 (1.18-1.23) | 3.72E-56 | 1.25 (1.22-1.28) | 2.84E-79 |
| Immune2 | 1.19 (1.16-1.22) | 4.89E-49 | 1.25 (1.22-1.27) | 5.81E-77 |
| EC-Meso | 1.17 (1.14-1.2) | 3.95E-39 | 1.2 (1.17-1.23) | 8.93E-52 |
| EC | 1.17 (1.14-1.2) | 7.07E-39 | 1.19 (1.16-1.22) | 2.86E-49 |
| PC-SMC | 1.19 (1.16-1.22) | 8.49E-49 | 1.24 (1.22-1.27) | 1.53E-76 |

HR: hazard ratio; CI: confidence interval; aCM: atrial cardiomyocyte; vCM: ventricular cardiomyocyte; Adip: adipocyte; FB: fibroblast; NC: neuronal cell; EC: endothelial cell; Meso: mesothelial cell; PC: pericyte; SMC: smooth-muscle cell.

We further evaluated whether integrating csPRSs and their corresponding c-csPRSs derived from the cell-cluster training strategy could improve CAD risk prediction. Ridge regression, elastic net regression, and XGBoost models were trained using csPRSs and c-csPRSs as predictors and compared with the genome-wide CAD PRS generated using SDPR weights. In the UKB testing dataset, the genome-wide CAD PRS achieved a C-index of 0.59 (95% CI, 0.57-0.60). The integrated models showed similar predictive performance, with C-indices of 0.59 (95% CI, 0.58-0.61) for ridge regression, 0.59 (95% CI, 0.58-0.61) for elastic net regression, and 0.59 (95% CI, 0.58-0.60) for XGBoost, respectively. Although ridge and elastic net models showed marginal improvements, integrating csPRSs did not substantially increase prediction accuracy beyond that of the genome-wide CAD PRS.

### Associations with CAD and its risk factors

Among testing samples, all 10 csPRSs trained using the cell-cluster strategy were significantly associated with incident CAD in Cox proportional hazards models, with hazard ratios ranging from 1.14 (95% CI, 1.11-1.18) for the NC csPRS to 1.25 (95% CI, 1.22-1.29) for the Immune 2 csPRS (Figure 4). Compared with the genome-wide CAD PRS (HR, 1.40; 95% CI, 1.36-1.44), csPRSs showed attenuated effect sizes, likely reflecting the smaller number of variants included in each score, and disease heterogeneity.

**Figure 4.**
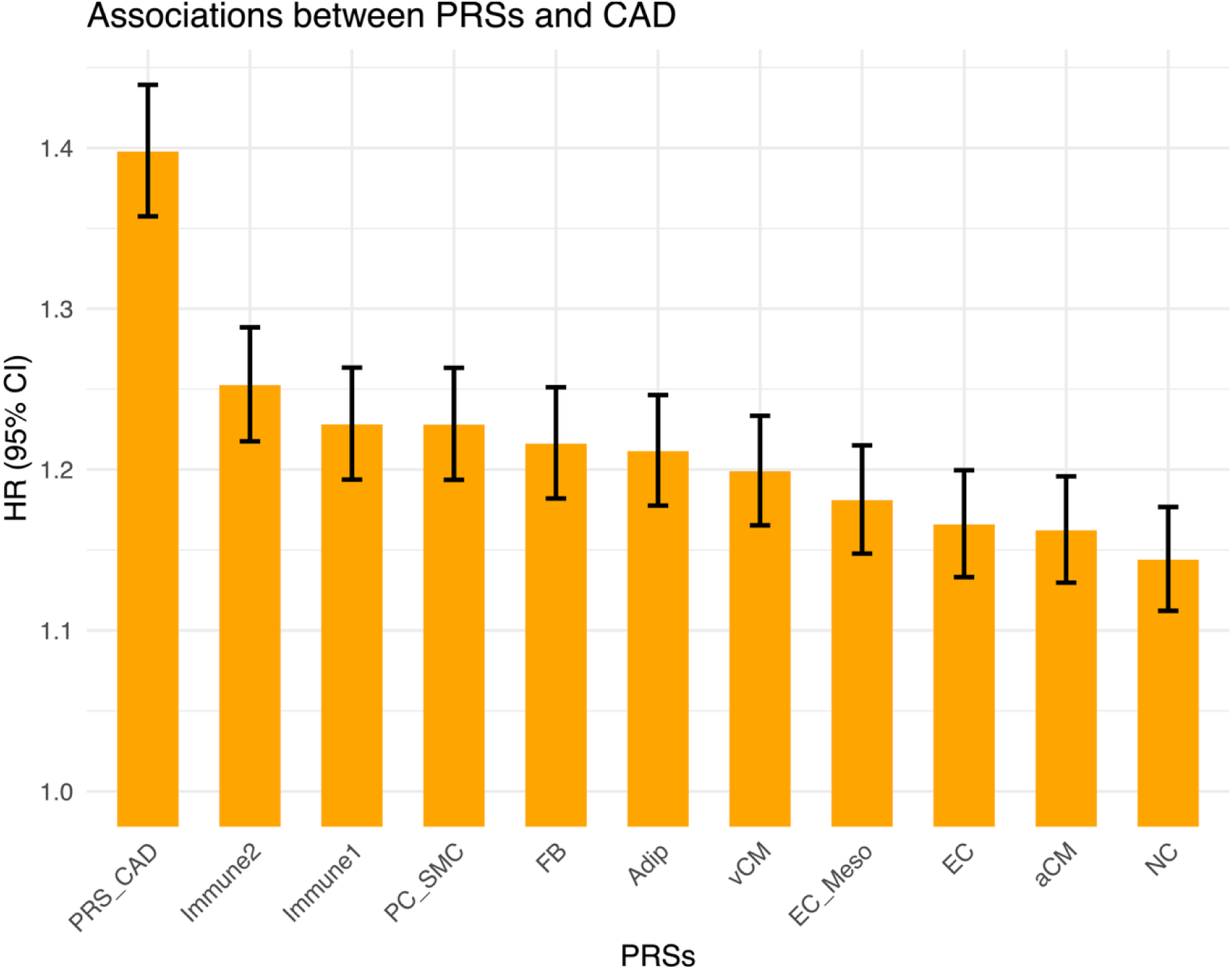
HRs of csPRSs and CAD PRS in testing samples. HRs and 95% CIs for incident CAD across CAD PRS and 10 csPRSs among testing samples. Our derived csPRSs showed significant associations with CAD while the magnitudes were lower than the genome-wide PRS.

We further evaluated associations between the 10 csPRSs, the genome-wide CAD PRS, and 71 phenotypes to characterize shared and distinct genetic architectures. Phenotypes associated with the genome-wide CAD PRS, such as lipid phenotypes, were generally also associated with multiple csPRSs, suggesting pervasive pleiotropy (Supplementary Figure 4). For example, the genome-wide CAD PRS showed strong associations with lipid phenotypes, including LDL-C (p-value = 1.06×10^-59^), and each csPRS was significantly associated with at least one lipid-related phenotype. However, the strength of associations varied substantially across csPRSs. Notably, the Adip csPRS demonstrated the strongest and broadest associations across lipid traits, being significantly associated with all lipid-related phenotypes examined. These findings suggest that csPRSs retained known pleiotropy associations while further localizing genetic effects to specific biologically relevant cell clusters, such as the enrichment of lipid-associated genetic signals within the Adip csPRS.

### Interactions between csPRSs and lipid phenotypes

Interaction effects on CAD between 11 PRSs (the genome-wide CAD PRS and 10 csPRSs) and 8 lipid-related phenotypes were evaluated using the testing dataset. Among the 88 pairs tested, 20 showed nominal significance (P<0.05) (Figure 5 and Supplementary Table 7). The genome-wide CAD PRS significantly interacted with lipid-lowering medication use, LDL-C, HDL-C, and ApoA to affect CAD risk. Six csPRSs also showed significant interactions with lipid-lowering medication use, with HRs ranging from 0.91 (95% CI, 0.85-0.96) for the PC-SMC csPRS to 0.94 (95% CI, 0.88-1.00) for the NC csPRS. Notably, the Adip and EC csPRSs showed significant interactions with total cholesterol, with HRs of 1.06 (95% CI, 1.03-1.10) and 1.03 (95% CI, 1.00-1.07), respectively. The Adip csPRS also showed positive interactions with LDL-C (HR, 1.09; 95% CI, 1.04-1.13) and ApoB (HR, 1.29; 95% CI, 1.12-1.49). In contrast, EC-Meso, FB, and Immune 1 csPRSs showed significant interactions with HDL-C and ApoA. These findings demonstrated distinct interaction patterns across cell-type-specific genetic components and suggested that the Adip and EC csPRSs captured biologically relevant lipid interactions not identified using the genome-wide CAD PRS alone.

**Figure 5.**
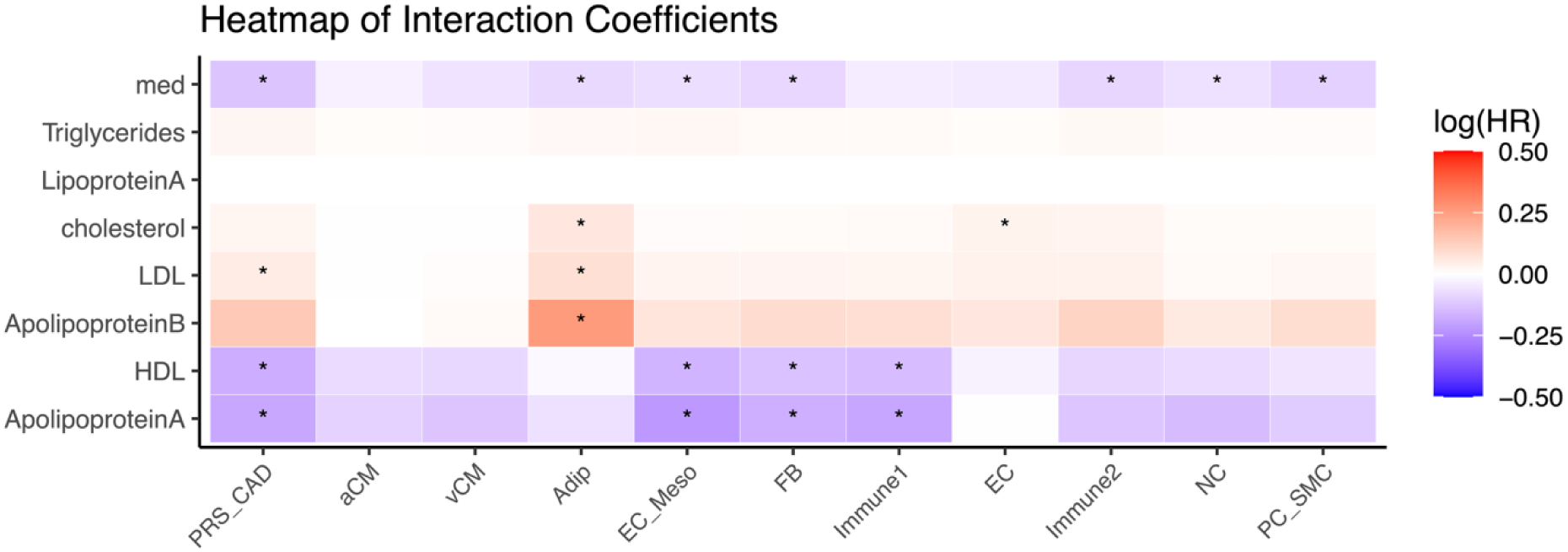
Interactions between PRSs and lipids-related phenotypes. Heatmap of beta coefficients for interactions between eleven PRSs and eight lipids-related phenotypes. Significant interactions with p-value < 0.05 were annotated with asterisk. Adip csPRS showed significant interactions with Apolipoprotein B, total cholesterol, LDL, and lipids-lowering medicine, while EC-Meso, FB, Immune1 csPRSs interacted with two good lipids: Apolipoprotein A and HDL.

We further evaluated multiplicative and absolute interactions using categorized PRSs and lipid-related phenotypes. P-values for multiplicative interactions and trends in ARR are summarized in Supplementary Table 8, and significant multiplicative interactions are presented in Figure 6. The genome-wide CAD PRS showed significant multiplicative interactions with HDL-C (P for interaction=2.35×10^-2^) and lipid-lowering medication (P for interaction=2.06×10^-3^). Similarly, EC-Meso and FB csPRSs interacted with HDL-C, whereas Adip, PC-SMC, and Immune2 csPRSs interacted with lipid-lowering medication, suggesting that these genome-wide interactions may be partially attributable to specific cell-type-related genetic components.

**Figure 6.**
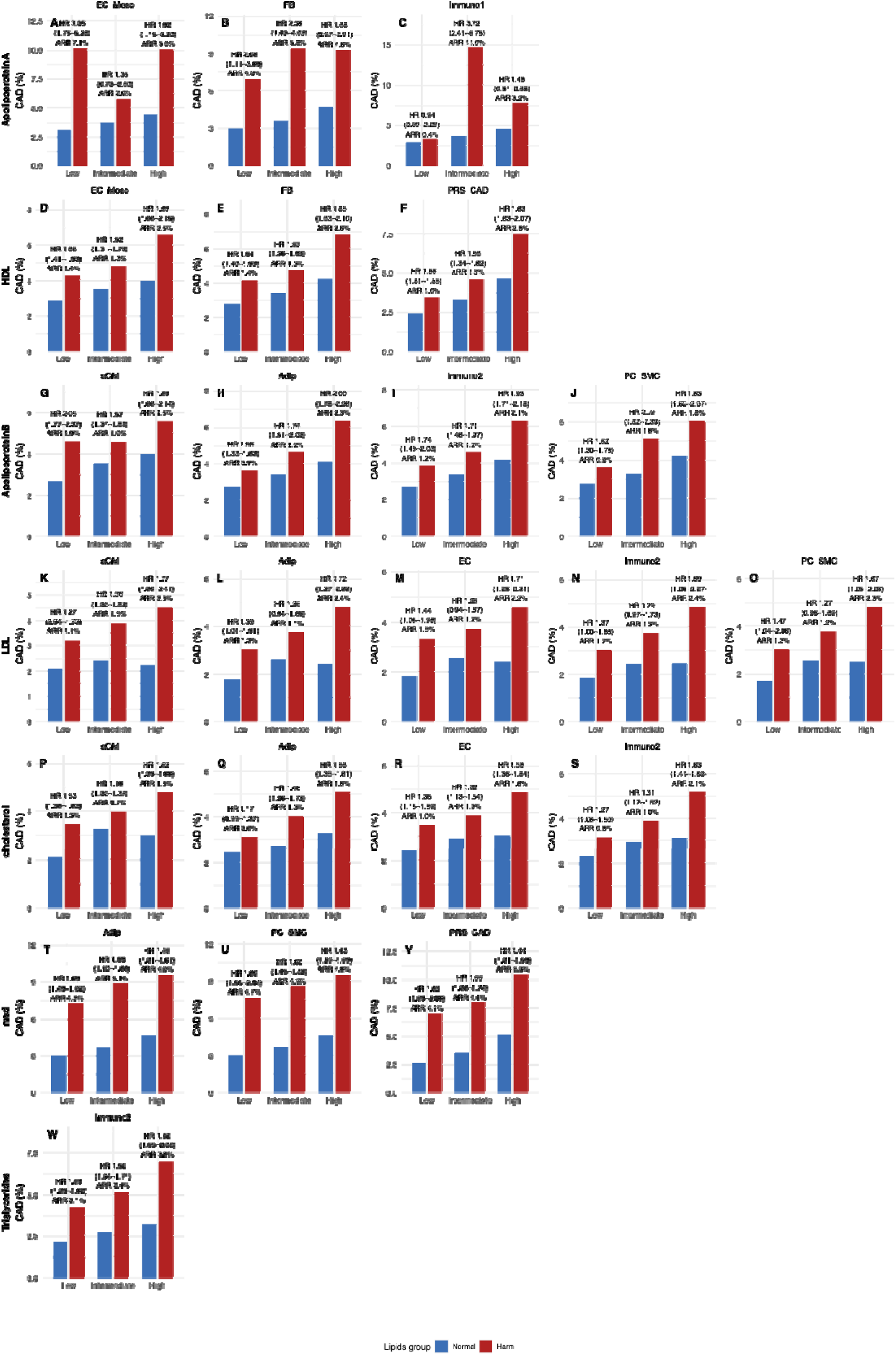
Interactions between binary lipids phenotypes and PRS tertiles. Bar plots for proportions of incident CAD cases in groups defined using binary lipids phenotypes and PRS tertiles for 23 lipids-PRS pairs with significant multiplicative interactions. ARRs between lipids groups were calculated. The associations between binary lipids phenotypes and CAD stratified by PRS tertiles were annotated above the ARRs. Each column contained results for one lipids phenotype, and each sub-figure was for one PRS. Compared to CAD PRS, csPRSs showed significant interactions with more lipids phenotypes. For example, Adip csPRS interacted with Apolipoprotein B, LDL, total cholesterol, and lipids-lowering medications.

In addition, at least one csPRS showed significant interaction with each lipid-related phenotype except LipoA, for which no significant interactions were observed. Notably, the Adip csPRS significantly interacted with ApoB (P for interaction=6.97×10^-3^), LDL-C (P for interaction=2.15×10^-3^), and cholesterol (P for interaction=2.49×10^-3^), suggesting that the effects of LDL-related lipids on CAD risk may be modified by adipocyte-related genetic susceptibility.

Although few absolute interactions reached statistical significance, participants within the highest tertile of the Adip csPRS consistently showed the largest ARR associated with harmful ApoB, LDL-C, and cholesterol levels (2.3%, 2.4%, and 1.8%, respectively) compared with intermediate and low tertiles, supporting clinically meaningful differences in absolute risk.

We further hypothesized that CAD patients in the highest csPRS tertiles might exhibit distinct comorbidity patterns before CAD onset. Using PERMANOVA on disease-embedding profiles, we identified significant heterogeneity for six csPRSs: PC-SMC, Immune1, Adip, Immune2, vCM, and NC (Supplementary Table 9). Among these csPRSs, the proportion of variance explained by differences in disease trajectories ranged from 0.31% for NC to 0.45% for PC-SMC. These findings suggested that individuals with elevated csPRSs may follow distinct pre-CAD comorbidity trajectories compared with those with lower csPRS levels.

### Validation of identified interactions

We further evaluated the 23 PRS-lipid pairs showing significant interactions in the internal validation dataset.

In the internal validation dataset, consisting of 1,660 incident CAD cases and 38,677 controls, four interactions remained significant: interactions between the Adip csPRS and ApoB, LDL-C, and total cholesterol, as well as the interaction between the PC-SMC csPRS and lipid-lowering medication use (Supplementary Table 10). Stratified associations between lipid phenotypes and CAD across csPRS tertiles are shown in Supplementary Figure 4. For the three interactions involving the Adip csPRS, increasing trends in both hazard ratios and absolute risk reductions were observed across tertiles. For example, the association between ApoB and CAD increased from 1.62 (95% CI, 1.23-2.12) in the lowest tertile of the Adip csPRS to 2.35 (95% CI, 1.91-2.88) in the highest tertile, while the corresponding ARR increased from 0.9% to 2.9%. These findings suggest consistent modification of LDL-related lipid effects by adipocyte-related genetic risk. In contrast, for the interaction between the PC-SMC csPRS and lipid-lowering medication use, hazard ratios showed decreasing trends across tertiles, whereas ARRs remained relatively stable, suggesting weaker differences in absolute risk.

### Comparison with PD-PRS and EC PRS

Previous studies identified two biologically informed components of CAD genetic risk, lipid-related PD-PRS and EC PRS, that may partially explain gene-lipid interactions. We therefore compared these previously established scores with the csPRSs developed in this study.

The lipid-related PD-PRS showed significant correlations with all 10 csPRSs, with the strongest correlation observed for the Adip csPRS (r=0.26; Supplementary Table 11), suggesting partial overlap in the biological mechanisms captured.

Of the previously reported EC PRS consisting of 35 variants, 9 overlapped with those included in our analyses (Supplementary Table 12). Notably, these variants were not exclusive to EC-related csPRSs; seven were included in the FB csPRS, indicating that variants assigned to EC-related functions may also contribute to biological processes in other cell types. Consistent with this observation, significant correlations between the EC PRS and csPRSs were observed across multiple clusters, ranging from 0.05 for the NC csPRS to 0.35 for the FB csPRS (Supplementary Table 11). The Adip csPRS showed a moderate correlation with the EC PRS (r=0.15), while the two endothelial-related csPRSs, EC and EC-Meso, showed somewhat stronger correlations (r=0.17 and r=0.31, respectively). These findings suggest that variants included in the EC PRS may capture functions spanning multiple cell types and partially overlap with adipocyte-related genetic components.

Overall, the Adip csPRS showed similarities to two previously established partial PRSs associated with lipid-related interactions in CAD.

## Discussion

In this study, we developed 10 csPRSs for CAD using variants mapped to genes specifically expressed in distinct cardiac and vascular cell populations. These csPRSs showed distinct interaction patterns with lipid-related phenotypes on CAD risk, highlighting biological heterogeneity underlying genome-wide CAD genetic risk. Among the identified interactions, the Adip csPRS showed the strongest and most consistent interactions with ApoB, LDL-C, and total cholesterol in the UKB. These findings suggest that adipocyte-related genetic susceptibility may modify the adverse effects of LDL-related lipids on CAD risk and the potential utility of csPRSs for identifying individuals who may be particularly vulnerable to hyperlipidemia-driven CAD.

We further compared two strategies for csPRS construction: genome-wide and cell-cluster-specific training. The csPRSs derived from the cell-cluster training strategy consistently showed stronger associations with CAD and modestly improved predictive performance compared with those derived from genome-wide training. These findings suggest that cell-type-specific gene expression information meaningfully contributes to genetic risk estimation for CAD, and that incorporating biologically informed annotations during PRS weight estimation may improve the specificity of genetic risk modeling. Our results are consistent with previous studies demonstrating the value of integrating scRNA-seq data with genetic analyses in CAD and other complex diseases^15,27–29^.

In addition, we identified distinct interaction patterns that were not captured by the genome-wide CAD PRS. LDL-related lipid traits (ApoB, LDL-C, and cholesterol) primarily interacted with the Adip and EC csPRSs, whereas HDL-related traits (ApoA and HDL-C) interacted with the EC-Meso, FB, and Immune1 csPRSs. These findings suggest that different lipid subclasses may influence CAD risk through distinct biological pathways. LDL-related lipids are directly involved in cholesterol deposition and plaque formation during atherosclerosis development, particularly through endothelial dysfunction and lipid accumulation within the arterial wall^30^. Also, our results suggested that the LDL-mediated CAD risk can involve adipocytes. In contrast, HDL-related lipids are thought to exert protective effects primarily through reverse cholesterol transport and modulation of vascular inflammation rather than directly reducing plaque burden^31^. Our results suggest the HDL function on CAD might involve certain subtypes of ECs, mesothelial cells, FB, and immune cells such as macrophages, reinforcing the close relationship with inflammation. Collectively, these results demonstrate that decomposing genome-wide PRSs into cell-type-specific components may reveal biologically meaningful interactions masked in conventional PRS analyses and provide additional insights into the heterogeneous mechanisms underlying CAD.

Of note, we identified and replicated the interactions between Adip csPRS and three LDL-related lipids: ApoB, LDL-C, and cholesterol. The Adip csPRS primarily contained genes expressed specifically in adipocytes, and these genes showed enrichment for lipid metabolism pathways such as the PPAR signaling pathway and the insulin resistance pathway Supplementary Table 4. Our results support the important role of adipocytes, or, more broadly, adipose tissue, in atherosclerosis, particularly regarding LDL-related lipids. Although adipocytes are not directly involved in plaque formation and rupture, adipose tissue is vital for whole-body metabolism, inflammation, and endocrine functions, and thus can promote LDL accumulation, thereby accelerating atherosclerosis^32^. Taken together, adipocytes can contribute to atherosclerosis, and the Adip csPRS we developed aggregated the genetic liability that interacted with LDL-related lipids and might be used to identify those who were more susceptible to CAD due to high LDL-related lipids.

We compared established PRSs that interacted with lipids—lipids-PD-PRS and EC PRS—with the csPRSs. The lipids-PD-PRS, constructed variants, located genomic regions where CAD showed local genetic correlations with a variety of lipids and captured the genetic liability to CAD that can be explained by pleiotropy with lipids^10^. This PD-PRS did not show strong positive interactions with all three LDL-related lipids as we observed for Adip csPRS, suggesting variants contributing to the interactions were excluded. The partial correlation with Adip csPRS supported the hypothesis. The EC PRS, comprising variants involved in endothelial cell functions, showed strong associations with LDL-C^11^. By comparing variants with those included in our csPRSs, we found that not all variants are specific to endothelial cells and can function in other cell types, such as adipocytes. Combining previous evidence and our results, we hypothesized that variants that modified associations between LDL-related lipids and CAD onset might be related to different cell types such as EC and adipocytes.

We note several limitations of our study. First, variants were mapped to genes based on genomic position using 10-kb upstream and downstream windows, which may both miss functionally relevant variants and include variants unrelated to gene regulation. In particular, regulatory variants identified through quantitative trait loci (QTLs) analyses, especially trans-expression QTLs, were not incorporated. Second, gene-expression patterns vary across tissues and physiological states. Our analyses were based on scRNA-seq data from heart tissue, which may not fully capture disease-relevant expression profiles in coronary arteries or atherosclerotic plaques. Another limitation is that genes included in the csPRSs were identified based on differential expression rather than causal evidence. Therefore, genes specifically expressed in certain cell types should not be interpreted as necessarily causal genes for CAD or lipid-related mechanisms. Third, although several interactions were replicated in the internal validation dataset, the extent to which these findings generalize beyond the UKB remains unclear. This is because interaction analyses require substantially larger sample sizes than marginal association for replication. We estimated that replication of an interaction with the observed effect size at a statistical significance of 6.97×10^-3^ would require about 115,340 participants, including approximately 5,147 incident CAD cases, to achieve 50% power. Therefore, substantial efforts are needed to replicate the results through population studies. Finally, all analyses were restricted to individuals of European ancestry, and the findings may not generalize to other ancestral populations.

## Conclusions

To sum up, we developed 10 cell-type-specific PRSs for CAD onset in this study and found distinct lipid-interacting patterns. We found that the Adip csPRS could modify associations between LDL-related lipids and CAD. By decomposing polygenic risk into cell-type-specific genetic components, we showed that CAD is likely driven by biologically distinct and lipid-dependent mechanisms that may be obscured by genome-wide PRS aggregation.

## Data availability statement

The individual genotype and phenotype data underlying this article were provided by the UK Biobank by permission (ref: 29900), and the instructions to apply for the data can be found at https://www.ukbiobank.ac.uk/enable-your-research/apply-for-access. The CAD GWAS summary statistics and scRNA-seq data were downloaded from publicly available databases, and the information on related articles was available in the Methods section. The summary-level data (e.g., PRS weights) will be available on the PGS catalog (https://www.pgscatalog.org/) once published.

## Author contributions

J.H., L.X., and H.Z. contributed to the study conception and design. Data requests and analysis were performed by J.H. and H.Z. T.L. and W.Z. helped with the embedding generation. The first draft of the manuscript was written by J.H. and H.Z., and all authors commented on the manuscript. All authors read and approved the final manuscript.

## Acknowledgements

We conducted the research using the UK Biobank resource under an approved data request (ref: 29900). We thank the GWAS consortium for making the GWAS summary data publicly accessible. We thank the authors of the scRNA-seq data for making the data publicly accessible. We thank the dbGaP for accessing the phenotype and genotype data for ARIC (accession numbers: phs001211.v5.p4 and phs000280.v9.p3). We also thank the Yale Center for Research Computing for using the McCleary High Performance Computing cluster. The funder had no role in the design of the study; the collection, analysis, or interpretation of the data; or the writing of the manuscript and decision to submit it for publication.

## Declaration of generative AI and AI-assisted technologies in the writing process

During the preparation of this work the authors used ChatGPT in order to improve writing. After using this tool/service, the authors reviewed and edited the content as needed and take full responsibility for the content of the publication.

## Ethics statement

The UK Biobank study had ethical approval from the United Kingdom National Health Service (NHS) Research Ethics Committee (Reference: 11/NW/0382). Signed and informed consent was obtained from all of the participants.

