## Supplementary figures for "Cell-type-specific polygenic risk scores reveal adipocyte-related interactions with lipids in coronary artery disease"

**Supplementary Figure 1. Proportion of variance in Jaccard similarity explained by clusters.**

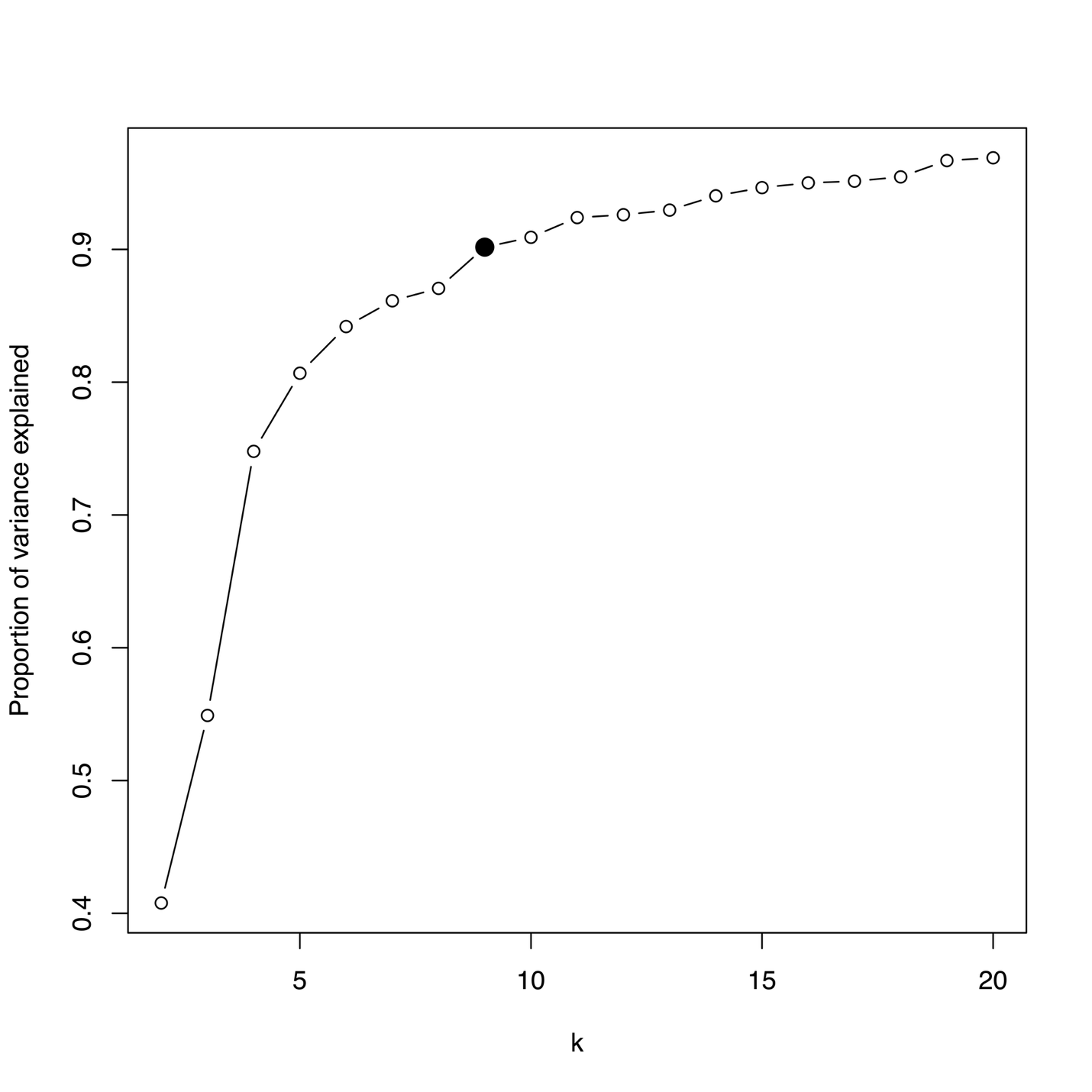

Proportion of variance in pairwise Jaccard similarity of gene lists for 13 major cell types and 64 subpopulations explained by clusters defined using hierarchical clustering. The y-axis is the proportion of variance explained, and the x-axis is the number of clusters (k) ranging from 2 to 20. The solid block point (k = 9) was selected with the cumulative variance explained more than 90%.

**Supplementary Figure 2. Heatmap for pairwise Jaccard similarity across 10 gene clusters.**

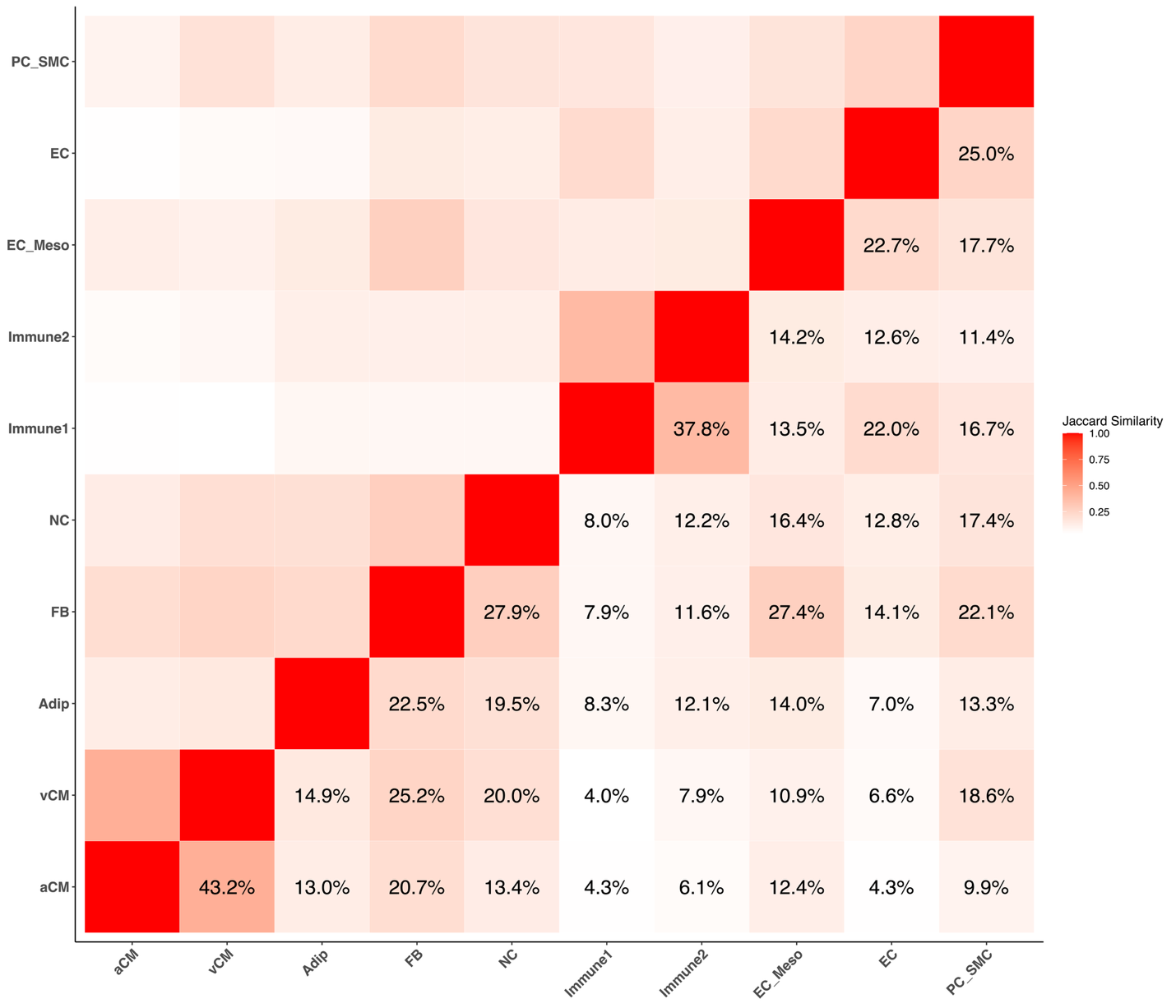

We calculated pairwise Jaccard similarity across 10 gene clusters. Each cell represented the Jaccard similarity between two clusters, and the numbers in the lower triangle were the Jaccard similarity. The similarities ranged from 4.0% for vCM and Immune 1 to 43.2% for aCM and vCM.

**Supplementary Figure 3. Associations between PRSs and 71 phenotypes.**

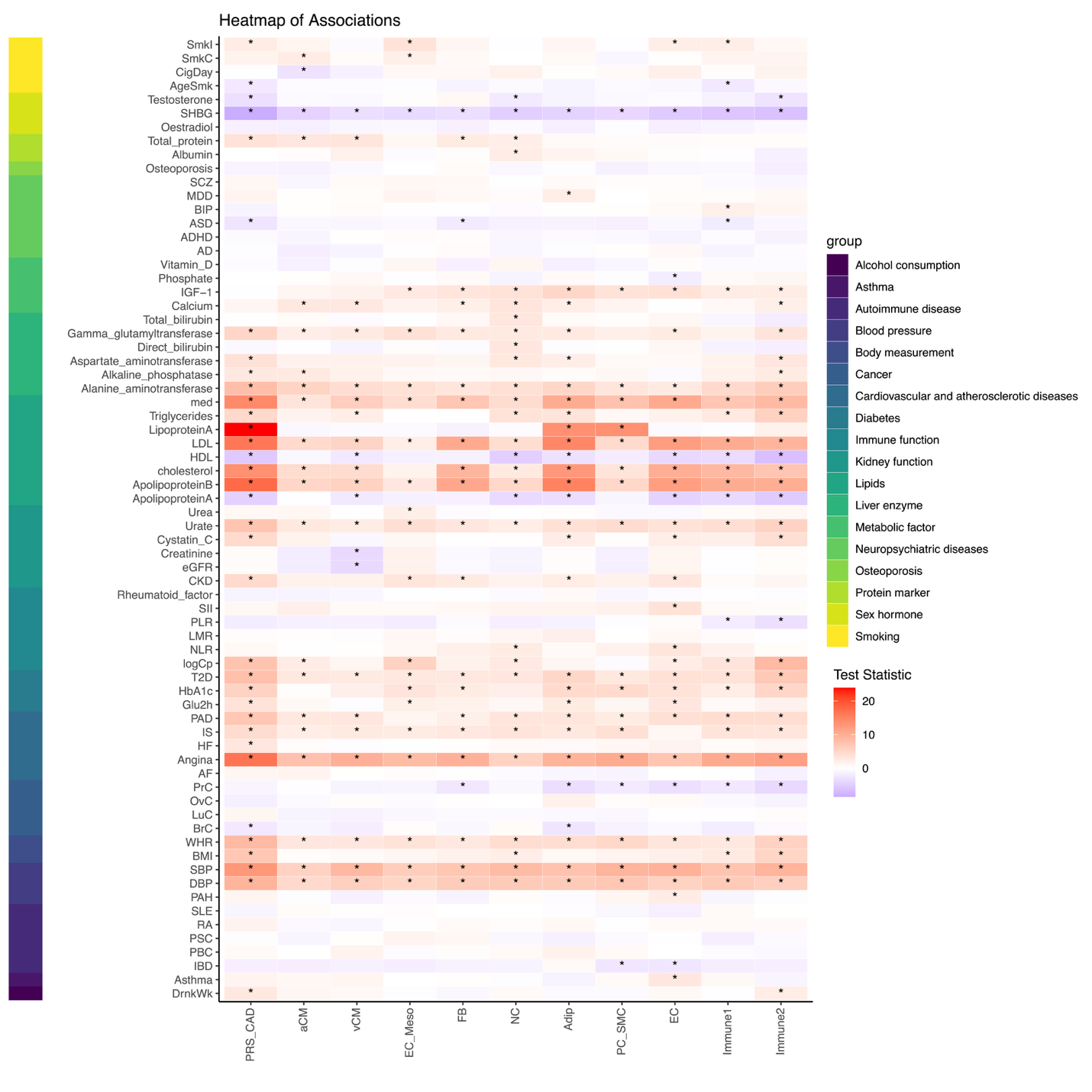

Heatmap for associations between 11 PRSs (CAD PRS and 10 csPRSs) and 71 phenotypes. Each cell represented the test statistic for each association, and significant associations (p-value < 0.05) were annotated with an asterisk. Phenotypes associated with CAD PRS were also associated with most of the csPRSs while the magnitude differed.

**Supplementary Figure 4.** **Interactions between binary lipid phenotypes and PRS tertiles in internal validation.**

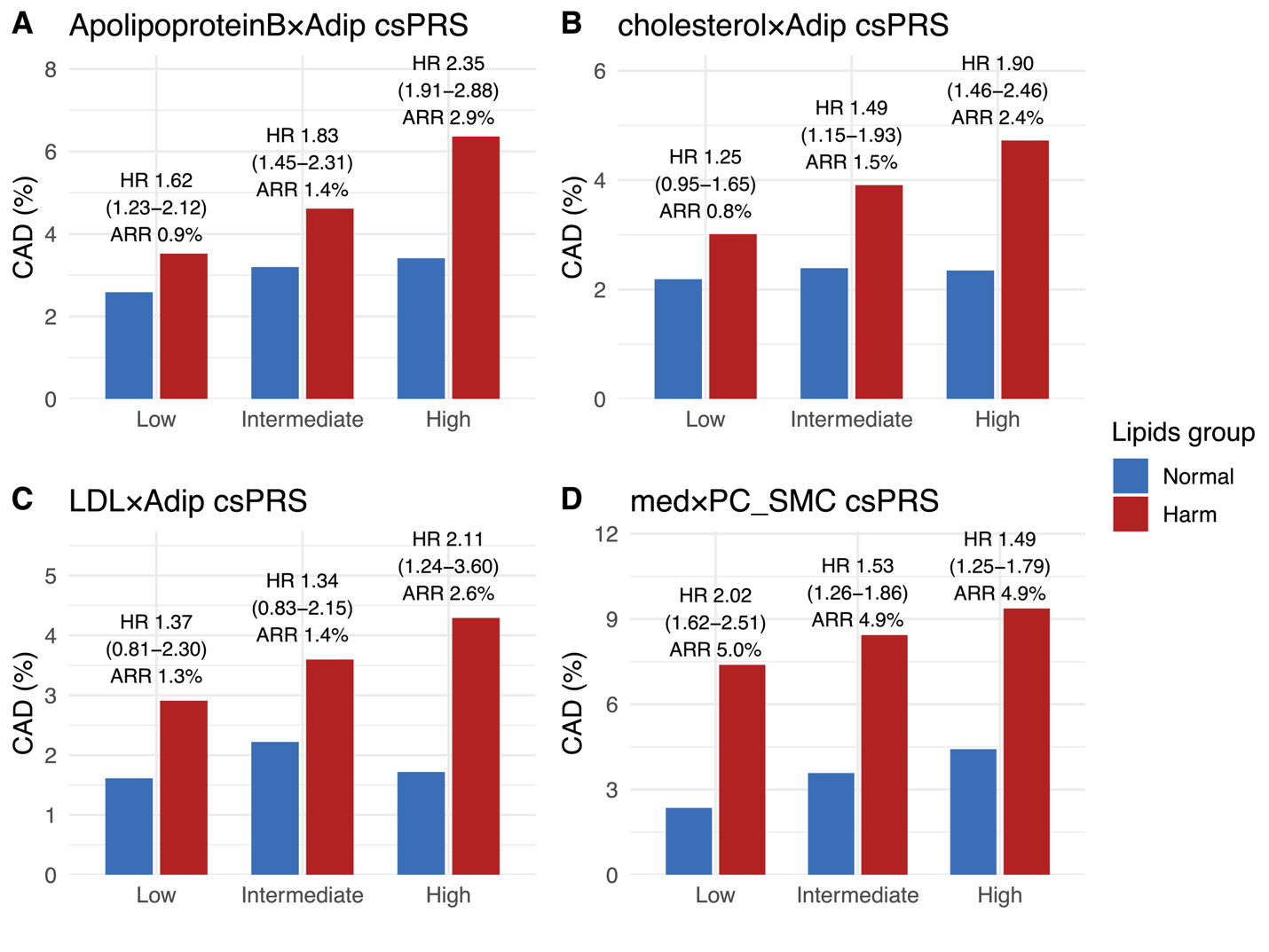

Bar plots for proportions of incident CAD cases in groups defined using binary lipid phenotypes and PRS tertiles for 4 lipid-PRS pairs with significant multiplicative interactions in internal validation. ARRs between lipids groups were calculated. The associations between binary lipids phenotypes and CAD stratified by PRS tertiles were annotated above the ARRs.
